# Characterization of SARS-CoV-2 Genetic Material in Wastewater

**DOI:** 10.1101/2021.07.19.21260777

**Authors:** Carolyn A Robinson, Hsin-Yeh Hsieh, Shu-Yu Hsu, Yang Wang, Braxton T Salcedo, Jessica Klutts, Sally Zemmer, Anthony Belenchia, Melissa Reynolds, Elizabeth Semkiw, Trevor Foley, XiuFeng Wan, Chris G. Wieberg, Jeff Wenzel, Chung-Ho Lin, Marc C Johnson

## Abstract

SARS-CoV-2 genetic material has been detected in raw wastewater around the world throughout the COVID-19 pandemic and has served as a useful tool for monitoring community levels of SARS-CoV-2 infections. SARS-CoV-2 genetic material is highly detectable in a patient’s feces and the household wastewater for several days before and after a positive COVID-19 qPCR test from throat or sputum samples. Here, we characterize genetic material collected from raw wastewater samples and determine recovery efficiency during a concentration process. We find that pasteurization of raw wastewater samples did not reduce SARS-CoV-2 signal if RNA is extracted immediately after pasteurization. On the contrary, we find that signal decreased by approximately half when RNA was extracted 24-36 hours post-pasteurization and ∼90% when freeze-thawed prior to concentration. As a matrix control, we use an engineered enveloped RNA virus. Surprisingly, after concentration, the recovery of SARS-CoV-2 signal is consistently higher than the recovery of the control virus leading us to question the nature of the SARS-CoV-2 genetic material detected in wastewater. We see no significant difference in signal after different 24-hour temperature changes; however, treatment with detergent decreases signal ∼100-fold. Furthermore, the density of the samples is comparable to enveloped retrovirus particles, yet, interestingly, when raw wastewater samples were used to inoculate cells, no cytopathic effects were seen indicating that wastewater samples do not contain infectious SARS-CoV-2. Together, this suggests that wastewater contains fully intact enveloped particles.

## 1 INTRODUCTION

Severe acute respiratory syndrome coronavirus 2 (SARS-CoV-2), the causative agent of coronavirus disease 2019 (COVID-19), was first identified in Wuhan, China in December 2019 and was declared a global pandemic by the World Health Organization (WHO) in March 2020. To date, SARS-CoV-2 has produced > 175 million cases and > 3.8 million COVID-19 related deaths worldwide (WHO, June 16^th^, 2021). SARS-CoV-2 has been shown to be spread primarily by respiratory droplets and occasionally by aerosols [1].

SARS-CoV-2 has 75-80% nucleotide similarity to severe acute respiratory syndrome coronavirus (SARS-CoV) that was responsible for outbreaks of severe acute respiratory syndrome in 2002 and 2003 in Guangdong Province, China[2-5]. Both SARS-CoV and SARS-CoV-2 use the cellular receptor Angiotensin-converting enzyme 2 (ACE2) which is highly expressed in the lung and oral mucosa and expressed at lower levels in the digestive tract [6-8]. Due partially to population sizes, material shortages, inaccessibility to laboratory equipment, a vast array of disease severity, and healthcare coverage concerns, it has not been possible to test every individual regularly for a SARS-CoV-2 infection. These disparities cause difficulty in monitoring community spread.

It has been reported that SARS-CoV, SARS-CoV-2, and other coronavirus RNA is detectable in feces of infected patients up to 6 to 10 days before symptom onset [9-14]. Additionally, screening of sewage for the prevalence of viruses including SARS-CoV-2, SARS-CoV, Poliovirus, *noroviruses, adenoviruses*, and *rotaviruses*, have been shown to correlate closely with the occurrence of cases in the community [15-20]. This supports wastewater surveillance as a useful method for monitoring community levels of SARS-CoV-2 infections.

Unlike from sputum, samples collected from feces of infected patients do not generally appear to contain infectious viral particles despite high levels of detectable viral RNA [21-23]; however, infectious particles cultured from feces has been reported before [24]. To date and to the best of our knowledge, there have been no confirmed cases of COVID-19 linked directly to wastewater treatment plants. Several groups have examined the survival rate of various other coronaviruses in raw, unpasteurized wastewater, and generally conclude that after a maximum of 3 days there is a 99.9% decrease in infectivity [25-27].

During periods of lower community infection rates, it was necessary to concentrate raw wastewater samples to reliably detect SARS-CoV-2 genetic material by quantitative reverse transcription polymerase chain reaction PCR (qPCR). Viral concentration from wastewater has been done using several different methods, and comparison of methods has been the focus of several manuscripts since early 2020 [28-30]. Our lab used a well-known Polyethylene glycol (PEG) and NaCl method for viral concentration that has been used for over 50 years [31-33]. As a control for viral recovery throughout concentration, we used a unique enveloped RNA virus. Interestingly, we noticed a disparity in recovery rates between our control virus and SARS-CoV-2 signal, leading us to question the nature of genetic material detectable in wastewater. In this manuscript, we examine recovery, temperature resistance, density, detergent resistance, and infectivity and conclude that the genomic material detected in wastewater is enveloped and non-infectious.

## 2 METHODS

### 2.1 Plasmids

The NL4-3 derived HIV containing the CMV driven Puromycin resistance gene and lacking the accessory genes Vif, Vpr, Nef, and Env was using InFusion Cloning (TaKara). To make this construct, we used a previously described NL4-3 derived HIV-CMV-GFP provided by Vineet Kewal Rammani (National Cancer Institute (NCI) – Frederick) [34]. This proviral vector lacks the accessory genes *vif, vpr, nef*, and *env* and contains a CMV promoter driven GFP in the place of *nef*. The NL4-3 derived HIV-CMV-GFP was digested using Stu1 and Xma1 (New England Biolabs (NEB)) to remove the GFP gene, and a gBlock fragment (Integrated DNA Technologies (IDT)) of a uniquely codon optimized Puromycin resistance gene was put in its place. The unique puromycin resistance gene sequence is as follows:

> ATGACAGAGTATAAGCCAACCGTCCGGCTCGCAACGAGAGACGATGTCCCGAGGGCAGTGCGCACGCTCGCC GCGGCCTTTGCGGACTACCCTGCAACAAGACACACTGTGGATCCCGATCGCCACATAGAGCGCGTGACTGAG CTGCAAGAACTGTTCCTTACCAGGGTGGGTCTCGATATCGGTAAGGTTTGGGTCGCCGACGACGGAGCGGCA GTGGCAGTCTGGACCACTCCTGAGAGCGTAGAAGCAGGCGCAGTGTTTGCAGAAATTGGCCCTAGAATGGCC GAATTGTCCGGTAGCCGGCTCGCTGCTCAGCAGCAGATGGAAGGCCTGCTCGCACCTCACAGACCCAAAGAA CCCGCGTGGTTCCTGGCGACAGTGGGAGTCAGTCCAGACCATCAGGGCAAAGGTCTCGGCTCAGCAGTTGTA CTGCCTGGGGTAGAGGCCGCAGAAAGGGCAGGGGTGCCGGCCTTCCTGGAAACATCTGCACCCAGAAACTT GCCTTTCTACGAGAGGCTGGGATTCACCGTTACCGCCGACGTGGAGGTGCCCGAAGGACCGCGCACTTGGTG CATGACGAGAAAGCCCGGGGCTTGA

To create the control sequence for qPCR standard curve, the plasmid described above was digested with EcoR1. A Wuhan strain N template was PCR amplified and infused using an InFusion kit (Takara).

### 2.2 Quantitative RT-qPCR assay

The TaqMan probe (VIC-5’ CGGTAAGGTTTGGGTCGCCGAC 3’-QSY) and the primer pair (*puro* Forward: 5’ CCCGATCGCCACATAGAGC 3’; *puro* Reverse: 5’ CCATTCTAGGGCCAATTTCTGC 3’) were designed and used to target the *puro* RNA described above. Primers and probe specificity was tested by BLAST analysis (NCBI) to prevent known nonspecific binding targets that could be obtained in a human specimen. The choice of VIC fluorescent dye for the puro TaqMan probe is for the application in the multiplex reactions with the SARS-CoV-2 N gene TaqMan probe utilizing the FAM reporter.

A plasmid (described above) carrying a unique *puro* resistance gene fragment along with a N gene fragment was constructed, purified from *Escherichia coli*, and used as standards for the RT-qPCR assay to ensure an equal molar ratio of puro and N gene detection. A standard curve was constructed at concentrations of 200,000 through 2 gene copies µL^-1^ and utilized to determine the copy number of the target *puro* gene in the spiked wastewater samples.

Final RT-qPCR one step mixtures consisted of 5 µL TaqPath 1-step RT-qPCR Master Mix (Thermo Fisher cat# A15299), 500 nM of each primer, 125 nM of each TaqMan probe, 5 µl of wastewater RNA extract and RNase/DNase-free water to reach a final volume of 20 µL. All RT-qPCR assays were performed in duplicate using a 7500 Fast real-time qPCR machine (Applied Biosystems). The reactions were initiated with 1 cycle of Uracil N-glycosylase (UNG) incubation at 25°C for 2 min to eliminate carryover and then 1 cycle of reverse transcription at 50°C for 15 min, followed by 1 cycle of activation of DNA polymerase at 95°C for 2 min and then 45 cycles of 95°C for 3 sec for DNA denaturation and 55°C for 30 sec for anneal and extension. The data is collected at the 55°C extension step.

### 2.3 Concentration and Recovery

Duplicate samples containing 50 mL of raw wastewater were collected and stored at 4°C. NL4-3 derived HIV containing the CMV driven Puromycin resistance gene and lacking the accessory genes Vif, Vpr, Nef, and Env were added to raw samples at a concentration of 4.8×10^7^ viral particles per sample. Raw samples were spun at 2,000xg for 5 minutes to remove large particulates, then vacuum filtered through a 0.22 µm filter (Millipore cat# SCGPOO525), mixed with a 50% (wt/v) Polyethylene glycol (PEG)(Research Products International (RPI) cat# P48080) and 1.2 M NaCl solution for a final concentration of 12% PEG and 0.3 M NaCl. Samples were mixed thoroughly and kept at 4°C for 1 hour, then spun at 12,000xg at 4°C for 2 hours. The pellet was extracted for RNA purification. RNA was extracted from the samples using the Qiagen QIAmp Viral RNA mini kit (cat# 52906) in a QIAcube Connect (Qiagen cat# 9002864). Additionally, 140 µL of wastewater was collected prior to filtering the sample, after filtration (before addition of PEG solution), and after concentration of virus. Viral recovery was determined by qPCR as described above.

### 2.4 Stability Assessment

Samples containing 50 mL of raw wastewater were collected and stored at 4°C. Samples were mixed gently and split into 3 × 16.7 mL aliquots. Aliquots were stored at either 4°C, Room Temperature (RT) (∼22 °C), or 37°C for 24 hours. After 24 hours, 140 µL of sample was collected, and RNA was extracted using the Qiagen QIAmp Viral RNA mini kit (cat# 52906) in a QIAcube Connect (Qiagen cat# 9002864). Viral recovery was determined by qPCR as described above. Statistics are a paired student’s t-test run on Microsoft Excel.

### 2.5 Pasteurization

Duplicate samples containing 50 mL of raw wastewater were collected and stored at 4°C. One 50 mL tube of each duplicate sample was kept at 4°C, and the other 50 mL tube of sample was incubated at 60°C for 2 hours then concentrated as described above. RNA was extracted from both pellets using the Qiagen QIAmp Viral RNA mini kit (cat# 52906) in a QIAcube Connect (Qiagen cat# 9002864). Viral recovery was determined by qPCR as described above.

For pasteurization effect on signal 24 hours later, duplicate samples were pasteurized and RNA extraction was done either immediately after the 2-hour incubation at 60°C or 24-36 hours after pasteurization. RNA was extracted from the samples using the Qiagen QIAmp Viral RNA mini kit (cat# 52906) in a QIAcube Connect (Qiagen cat# 9002864). Viral recovery was determined by qPCR as described above. Statistics are a paired student’s t-test run on Microsoft Excel.

### 2.6 Freeze-Thaw Sensitivity

Duplicate samples containing 50 mL of raw wastewater less than 1 week old were collected and stored at 4°C. One 50 mL tube of each duplicate sample was kept at 4°C, and the other 50 mL tube of sample was stored at -80°C. for 36 hours then thawed and concentrated as described above. RNA was extracted from both pellets using the Qiagen QIAmp Viral RNA mini kit (cat# 52906) in a QIAcube Connect (Qiagen cat# 9002864). Viral recovery was determined by qPCR as described above. Statistics are a paired student’s t-test run on Microsoft Excel.

### 2.7 Detergent Sensitivity

Duplicate samples containing 50 mL of raw wastewater were collected and stored at 4°C. One of each duplicate sample was treated with either 1% Triton X 100 or PBS for 2 hours at 37°C. RNA was extracted using the Qiagen QIAmp Viral RNA mini kit (cat# 52906) in a QIAcube Connect (Qiagen cat# 9002864). Viral recovery was determined by qPCR as described above. Statistics are a paired student’s t-test run on Microsoft Excel.

### 2.8 Density

Samples containing 50 mL of raw wastewater were collected and stored at 4°C. NL4-3 derived HIV containing CMV driven Puromycin resistance and lacking the accessory genes Vif, Vpr, Nef, and Env were added to raw samples at a concentration of 4.8×10^7^ viral particles per sample. Samples were concentrated as described above. Concentrated samples were then added to a density gradient ranging from 0% to 28% iodixanol in a 0.25 M sucrose dilutant according to the Optiprep protocol (Sigma cat# 92339-11-2). Gradients were spun in a Sorvall Discovery 100SE ultracentrifuge at 31,000 RPM for 3 hours at 4°C. After centrifugation, the gradient was fractioned. RNA was extracted from each fraction using the Qiagen QIAmp Viral RNA mini kit (cat# 52906) in a QIAcube Connect (Qiagen cat# 9002864). Viral recovery was determined by qPCR as described above.

### 2.9 Infectivity

Fresh samples were filtered through a 0.22 µm filter (Millipore cat# SCGPOO525). For the first recovery, 200 µl of each sample was inoculated to Vero E6 cells (CRL1586(tm), ATCC) in 6-well plates at a confluence of ∼90%. After one hour of adsorption, the inoculum was removed, and the cells were washed with PBS and covered with fresh optiMEM (Gibco, Thermo Fisher Scientific). Three days post inoculation, 1 mL of the supernatant from the last virus recovery was centrifuged and inoculated to fresh Vero E6 cells for the second and third virus recovery and cytopathic effect was observed. The SARS-CoV-2/human/USA/20×1003/2020 (GenBank Accession ID: MW521470.1) virus was used as the positive control at a multiplicity of infection (MOI) of 0.001.

## 3 RESULTS

### 3.1 SARS-CoV-2 Signal Recovery Through Concentration

Generally, it has been necessary to concentrate wastewater samples for reliable detection of SARS-CoV-2 by qPCR. Because concentration is sometimes necessary, it is important to know the rate of recovery throughout the process. During a period of higher community spread of COVID-19, we were able to reliably detect signal in raw, unconcentrated wastewater. This allowed us to compare signal before concentration and after each subsequent step (Figure 1). We extracted RNA from samples before filtration, after filtration, and after concentration for qPCR quantification. Unconcentrated sample numbers were multiplied based on the volume of original sample to ensure that recovery could be compared throughout the concentration process. Filtering preserved an average of 68% of signal and concentration preserved an average of 62% signal when compared to raw samples.

**Figure 1.**
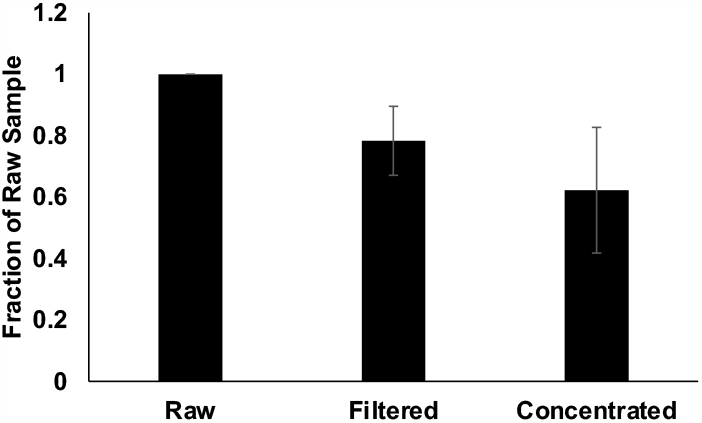
Recovery. N=9. Raw samples were spun at 2,000xg for 5 minutes to remove large particulates, then vacuum filtered through a 0.22 µm filter, and mixed with Polyethylene glycol (PEG) and NaCl solution for a final concentration of 12% PEG and 0.3 M NaCl. Samples were mixed thoroughly and kept at 4°C for 1 hour, then spun at 12,000 xg at 4°C for 2 hours. RNA was extracted from pellet, and viral recovery was determined by qPCR. Wastewater was collected prior to filtering the sample, after filtration (before addition of PEG solution), and after concentration of virus. Signal from unconcentrated samples was multiplied based on the total volume of sample to be concentrated to allow for equal comparison at each step. Error bars represent standard deviation.

### 3.2 Pasteurization and Freeze-Thaw Can Reduce SARS-CoV-2 Signal

Although wastewater is not thought to contain infectious SARS-CoV-2, raw wastewater contains a variety of other pathogens. Filtration through a 0.22 µm filter should remove many of these pathogens, but some have suggested pasteurizing samples at 60°C for 1 to 2 hours to inactivate potential pathogens [35]. To test the effect on signal due to pasteurization, duplicate samples were kept at either 4°C or 60°C for 2 hours prior to RNA extraction (Figure 2A). We found no significant difference between pasteurized and non-pasteurized samples (P value= 0.23) when RNA was extracted immediately after pasteurization; important to note that signal dropped significantly if samples were returned to 4°C after pasteurization and RNA was collected 24-36 hours after pasteurization was completed (Figure 2B). A 24–36-hour period between pasteurization resulted in a 44% reduction in signal (P value = 2.05×10^−6^). This finding is important as protocols using pasteurization as a method to inactivate pathogens in wastewater prior to RNA extraction and analysis may be time sensitive. Additionally, when samples were frozen at -80 °C for 36 hours and thawed prior to concentration, over 90% of the SARS-CoV-2 signal was lost (Figure 2C). This finding further stresses that processing of samples is time-sensitive, and storage of samples in a -80 °C will significantly disrupt signal.

**Figure 2:**
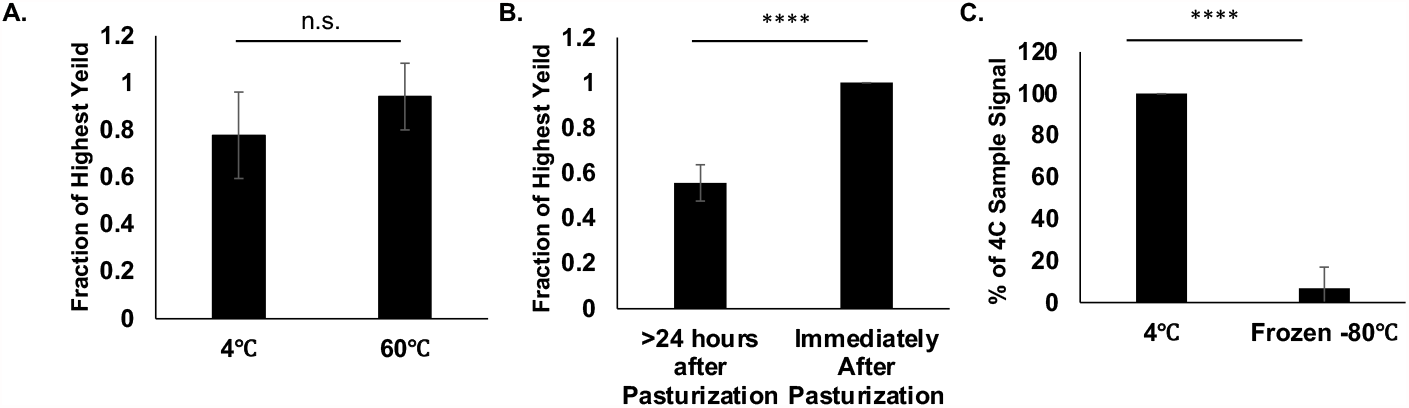
Impact of Pasteurization and Freeze-Thaw. **A)** N=6. Duplicate samples were kept at 4°C or incubated at 60°C for 2 hours. Raw samples were concentrated, and RNA was extracted from the pellet. Viral recovery was determined by qPCR. Fraction of Highest yield was calculated by the ratio signal between treatments. Error bars represent standard deviation. P-Value= 0.23 **B)** N=6. Duplicate samples were kept at 4°C or incubated at 60°C for 2 hours. RNA was extracted from raw samples either immediately after 2-hour incubation at 60°C or 24-36 hours after pasteurization. Viral recovery was determined by qPCR. Fraction of Highest yield was calculated by the ratio signal between treatments. Error bars represent standard deviation. P-Value= 2.05E-6. **C)** N=6. Duplicate raw samples were kept at kept at 4°C or -80 °C for 48 hours. Raw Samples were concentrated, and RNA was extracted from the pellet. Viral recovery was determined by qPCR. Error bars represent standard deviation. P-Value= 3.13E-6.

### 3.3 SARS-CoV-2 Signal is Higher than Enveloped Virus Control

As a control throughout wastewater screening, an NL4-3 derived HIV virus containing a CMV driven Puromycin resistance gene and lacking the accessory genes Vif, Vpr, Nef, and Env (Henceforth called ‘Puro Virus’) was added to raw wastewater samples at a concentration of 4.8×107 viral particles per sample (Figure 3A). Importantly, the Puro Virus contains a uniquely codon optimized puromycin resistance gene. The unique sequence present in the Puro Virus ensures that any signal detected throughout our experiments with this probe is from our internal matrix control. Both SARS-CoV-2 and the Puro Virus are positive-sense RNA contained in an envelope. Interestingly, upon comparison of signal detected in highly potent samples before and after concentration, SARS-CoV-2 recovery was consistently higher than the Puro Virus recovery (Figure 3B). On average, SARS-CoV-2 recovery was 2.7-fold higher than Puro Virus recovery. The consistent disparity between the SARS-CoV-2 recovery and the Puro Virus recovery was surprising and led us to hypothesize that the SARS-CoV-2 signal was coming from a different source than enveloped RNA, such as non-enveloped ribonuclear complexes, as we would expect that most enveloped particles would interact similarly with PEG during concentration. Following this finding, we aimed to characterize the SARS-CoV-2 genetic material detected in wastewater.

**Figure 3:**
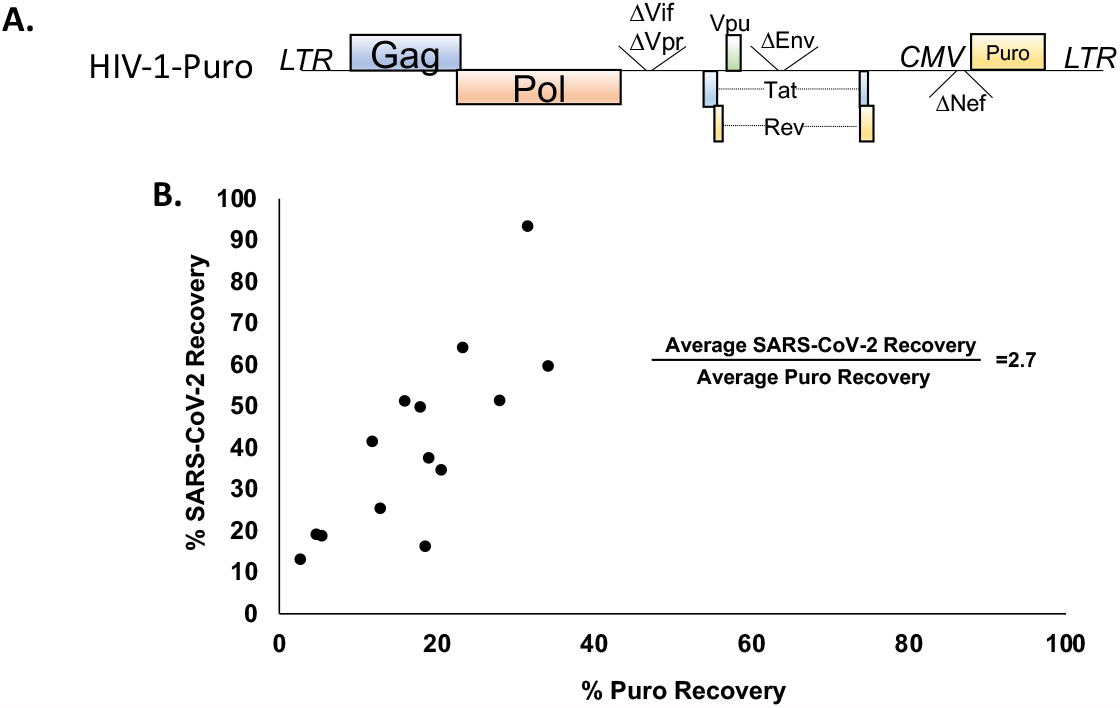
A) Schematic of Puro Virus Control. NL4-3 derived HIV containing CMV driven, uniquely codon optimized, Puromycin resistance and lacking the accessory genes Vif, Vpr, Nef, and Env. **B) Relative Recovery of Puro Virus Signal and SARS-CoV-2 Signal**. Samples were spiked with Puro Virus at a concentration of 4.8×107 viral particles per sample. RNA was extracted from samples both before and after concentration, and viral recovery was determined by qPCR.

### 3.4 SARS-CoV-2 Signal Stability

After collection from a sewer shed, samples are stored at 4°C until they are processed; however, we wanted to test the stability of samples at a variety of temperatures to determine if temperature control made a large impact on SARS-CoV-2 signal. A portion of each sample was kept at either 4°C, RT, or 37°C for 24 hours.

RNA was extracted from each sample, and SARS-CoV-2 signal was quantified using qPCR (Figure 4). Although samples that were maintained at 4°C had the least variability, there was no significant difference in signal from samples kept at any temperature (P values range 0.097 to 0.363). This finding is interesting as an increase in temperature from 4°C to either RT or 37°C could impact the activity of various enzymes that could be present in raw wastewater and impact rates of degradation of genetic material.

**Figure 4:**
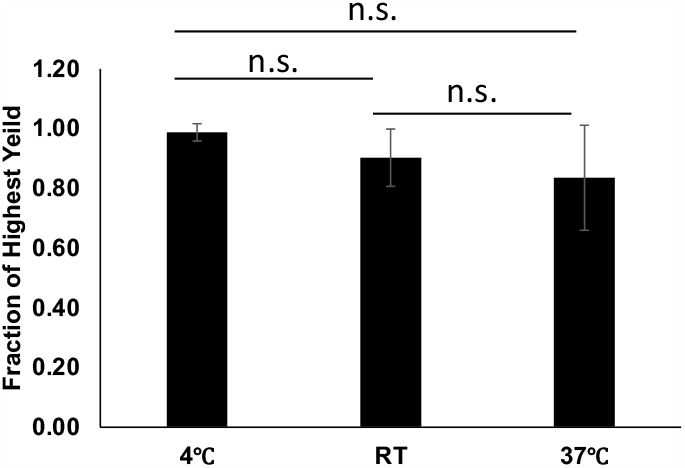
Temperature Stability. N=6. Samples were mixed gently and split into 3 aliquots. Aliquots were stored at either 4°C, RT, or 37°C for 24 hours. After 24 hours, RNA was extracted, and viral recovery was determined by qPCR. Error bars represent standard deviation. P-Values: 4°C to RT = 0.097, 4°C to 37°C = 0.108, RT to 37°C = 0.363.

### 3.5 Detergent Removes SARS-CoV-2 Signal

Because SARS-CoV-2 is an enveloped virus, it is very likely to be sensitive to detergent because of a disruption of the lipids composing the envelope. Presence of an envelope may protect genomic material from enzymes that may quickly degrade vulnerable genetic material. We were curious as to whether the SARS-CoV-2 signal detected in wastewater was sensitive to detergent. To answer this question, duplicate samples of raw wastewater were treated with either 1% TritonX-100 or PBS and kept at 37°C for 2 hours (Figure 5). RNA was extracted and samples were quantified using qPCR. Treatment with TritionX-100 reduced signal about 100-fold in comparison to samples treated with PBS alone. Contrary to our original hypothesis, this indicates that SARS-CoV-2 signal detected in wastewater is likely protected by a lipid bilayer, but this finding could also have other causes.

**Figure 5:**
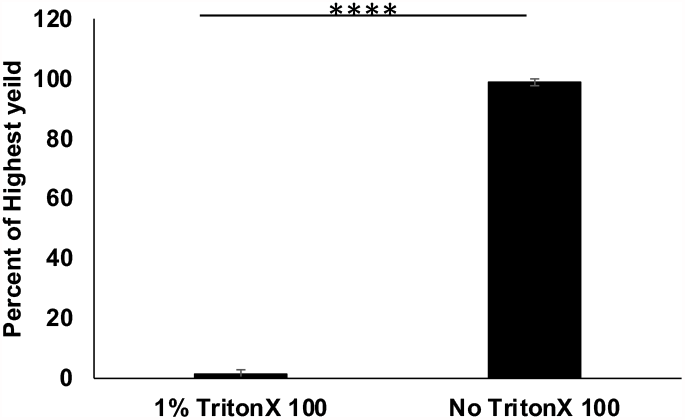
Detergent Sensitivity. N=6. Duplicate samples were treated with either 1% Triton X 100 or PBS for 2 hours at 37°C. RNA was extracted from raw samples, and viral recovery was determined by qPCR. Error bars represent standard deviation. P-Value= 2.10E-20.

### 3.6 Density of SARS-CoV-2

A loss of signal following treatment with detergent could be caused by a variety of things including breaking up of protein complexes. Because of this, we wanted to know if the density of SARS-CoV-2 particles was comparable to other known viral densities as a similar density would support that the particles are enveloped. To test density, concentrated wastewater samples also containing the Puro Virus at a concentration of 4.8×107 viral particles per sample were run through a density gradient containing 0% to 28% iodixanol in a 0.25 M sucrose dilutant and RNA was extracted from each fraction (Figure 6). Each fraction was then probed by qPCR for SARS-CoV-2 signal and the Puro Virus control. The fraction containing the highest signal for both the Puro Virus and SARS-CoV-2 correlates with a density (ρ) between 1.16 and 1.18 g.mL^-1^ as calculated according to the Optiprep protocol and confirmed by refractometer. This finding is in agreement with the expected density of retroviruses and further supports that the genetic material wastewater is similar to an enveloped viral particle [36].

**Figure 6:**
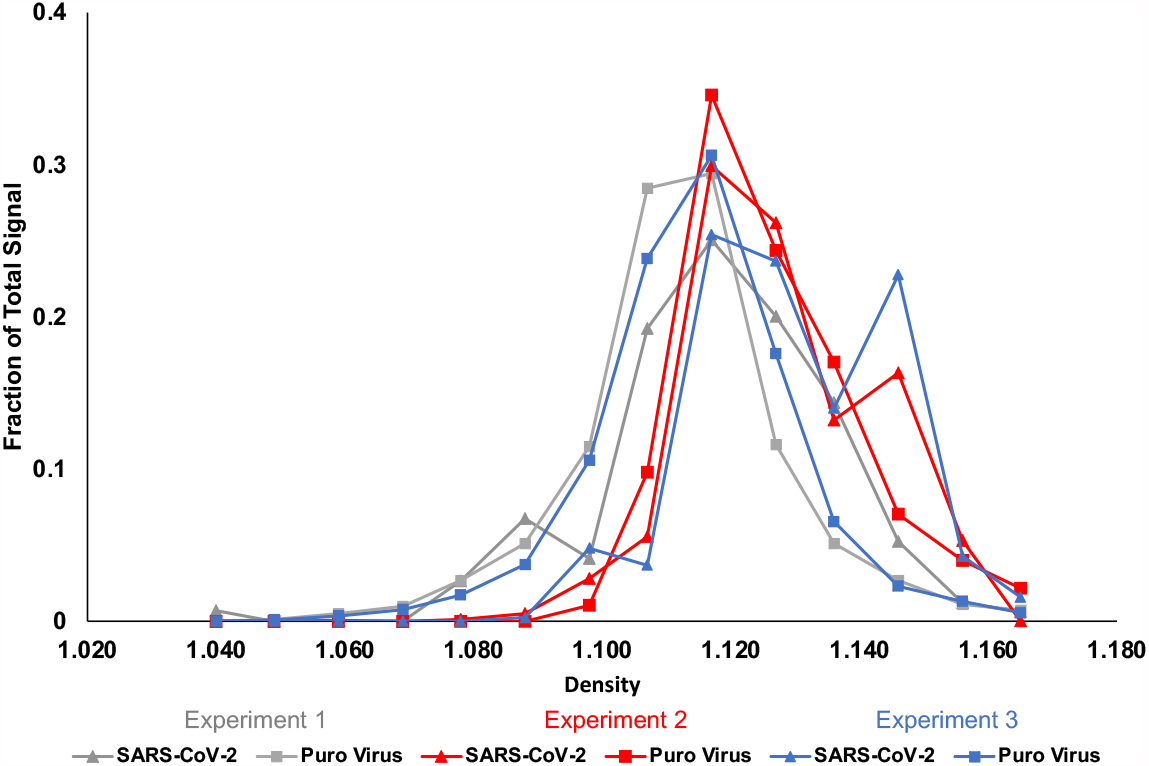
Density. N=3. Puro Virus was added to raw wastewater samples at a concentration of 4.8×10^7^ viral particles per sample. Raw samples were concentrated then added to a density gradient ranging from 0% to 28% iodixanol in a 0.25 M sucrose dilutant and spun in a Sorvall Discovery 100SE ultracentrifuge at 31,000 RPM for 3 hours at 4°C. RNA was extracted from fractions, and viral recovery was determined by qPCR.

### 3.7 No Cytopathic Effects from Wastewater Samples

The similarities in density along with detergent sensitivity suggest that the genetic material is enveloped. We were curious to know if wastewater samples contained infectious particles. To examine this question, aliquots of 10 raw wastewater samples from the week of June 28^th^, 2021, with SARS-CoV-2 signals ranging from 169,433 to 3,255,921 Copies/L were collected and used to inoculate Vero E6 cells within a week of collection. Seventy-two hours after inoculation, supernatant was collected and used to inoculate fresh Vero E6 cells. At each passage, cells were examined for cytopathic effects. No cytopathic effects were seen at any point during the experiment (Figure 7). The absence of cytopathic effects suggests that wastewater samples do not contain infectious SARS-CoV-2 Particles. As a positive control, cells were infected with SARS-CoV-2/human/USA/20×1003/2020 (GenBank Accession ID: MW521470.1) at a MOI of 0.001. These cells showed significant cytopathic effects whereas the negative control cells had no cytopathic effects.

**Figure 7:**
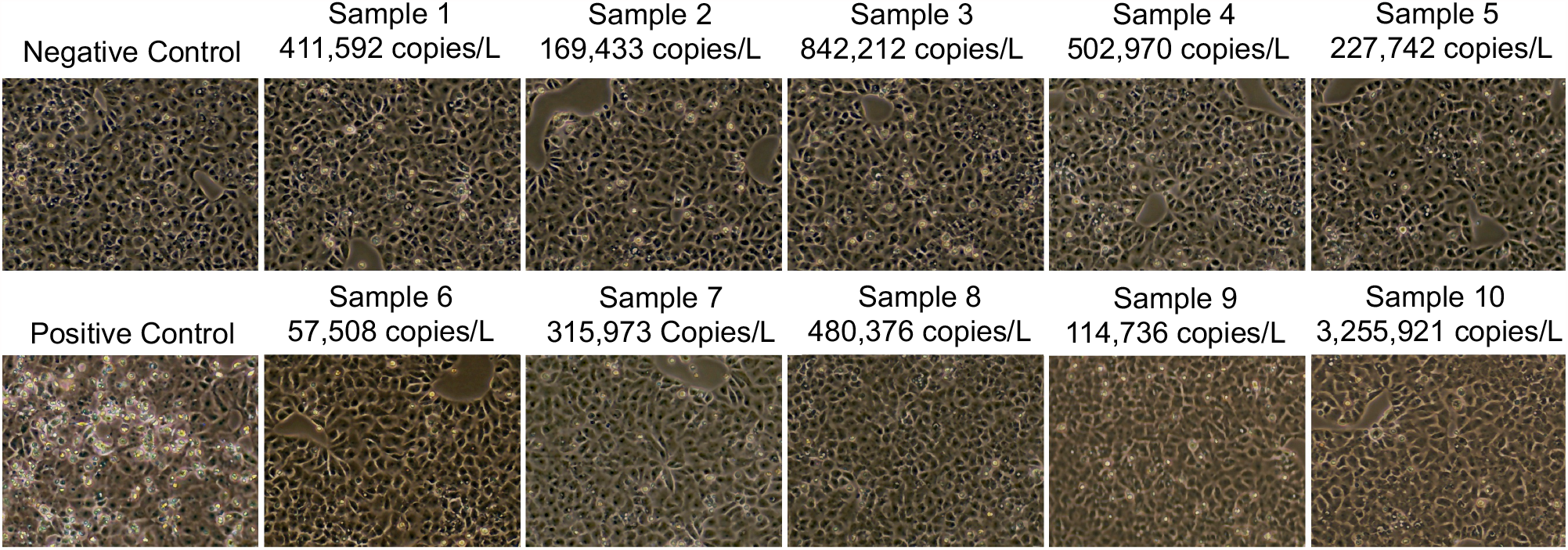
Infectivity. CPE of Vero E6 cells 3 days post inoculation with fresh wastewater samples. Images taken 3 days after the 3^rd^ inoculation. Ten total samples were tested. Numbers above each picture represent the copy number per liter from that sample as measured by qPCR.

## 4 DISCUSSION

COVID-19 causes a wide variety of symptoms and disease severity. This combined with inaccessibility to testing due partially to supply costs, availability, and varied access to healthcare makes accurately tracking cases of COVID-19 difficult. Like several other viruses, SARS-CoV-2 was shown to be present in the COVID-19 patient feces and therefore wastewater despite being primarily transmitted via respiratory droplets [9-13]. To date, there have been no confirmed cases of SARS-CoV-2 infection from wastewater treatment plants, and infectious virus has not been able to be reliably cultured from wastewater [25-27]. In early stages of the pandemic and during periods of lower community spread of SARS-CoV-2, it has been necessary to concentrate wastewater samples to reliably detect genetic material using qPCR. Using samples from communities with high community levels of COVID-19, we have shown that filtration and concentration of wastewater samples reliably reserves ∼60% of signal from raw wastewater, supporting that this method of filtering and concentration is a consistent method for concentrating and comparing community viral loads (Figure 1). While the method described here includes filtration through a 0.22 .m filter to remove other debris and bacterial pathogens, other groups have suggested pasteurizing wastewater samples to inactivate any pathogens present in wastewater. We have shown here that pasteurizing samples for 2 hours does not impact signal if RNA is extracted from samples immediately; however, this does not remain true for samples that have been pasteurized and returned to 4°C for RNA extraction between 24- and 36-hours post-pasteurization (Figure 2). While the decrease in signal is approximately 2-fold, it remains an important note that pasteurization may make a consistent experimental timeline of higher importance for those who are quantifying viral loads after pasteurization.

Interestingly, we noticed that recovery of the Puro Virus control was consistently about 3-fold lower than the recovery of SARS-CoV-2 through concentration (Figure 3B). Initially, this led us to believe that the SARS-CoV-2 signal present in wastewater was coming from a different source than an enveloped particle, but our data suggests the contrary and led us to investigate the nature of the genomic material producing signal.

From the time it is deposited into a sewer system to its arrival at a wastewater treatment plant, a fecal sample may go through a variety of temperature changes. Additionally, as temperatures get closer to 37°C, enzymes that degrade genetic material may become more active. We show that in a 24-hour period, temperature changes are tolerated as there is no significant difference in signals from samples kept at 4°C, room temperature, or 37°C (Figure 4). This finding is interesting as it suggests some sort of protection of genetic material from degradation and suggests that outdoor temperatures may not impact reliability of signal detection when levels of SARS-CoV-2 genetic material are monitored over time.

Because of high levels of genomic material found in feces, it was a high concern early in the pandemic that feces and wastewater may be a source infectious virus; however, many efforts to culture infectious virus from fecal or wastewater samples have failed [21-23]. Here we demonstrate that genomic RNA collected from wastewater samples is sensitive to detergent which suggests that the genomic material is protected by a lipid bilayer (Figure 5). Additionally, the material concentrated from wastewater samples has a very similar density to non-infectious retroviral particles again supporting that the material concentrated from wastewater shares similarities to an enveloped viral particle (Figure 6). In agreement with prior findings discussed above, wastewater samples did not contain any infectious SARS-CoV-2 particles (Figure 7). It is feasible that enzymes present in the digestive tract, such as Trypsin, cleave much of the Spike glycoprotein from virus present in the digestive tract resulting in viral particles that are enveloped yet not infectious. Further studies need to be done to determine whether the genomic material is full length SARS-CoV-2 genomic material and to investigate why the genomic material found in wastewater may be non-infectious while also remaining to be protected by a lipid bilayer.

## Data Availability

All data generated or analysed during this study are included in this published article.

## FUNDING ACKNOWLEGEMENT

Funding for the project was administered by the Missouri Department of Health and Senior Services (DHSS). This project was supported by funding from the Centers for Disease control and the National Institutes of Health grant U01DA053893-01

